# Non-negligible seroprevalence of murine typhus and its predictors in Japan: a large-scale seroepidemiological study

**DOI:** 10.1101/2023.01.12.23284497

**Authors:** Tetsuro Aita, Eiichiro Sando, Shungo Katoh, Sugihiro Hamaguchi, Hiromi Fujita, Noriaki Kurita

## Abstract

Scrub typhus (ST) and Japanese spotted fever (JSF) are endemic rickettsioses in Japan, whereas murine typhus (MT) has been slightly reported over the last 50 years. To elucidate the epidemiology and risk factors of MT, we conducted a cross-sectional study of residents in the southern Boso Peninsula, a rickettsia-endemic region, from August to November 2020, using their sera, questionnaires, residential addresses, and geographic information. A total of 2,382 residents were included in the study. The seroprevalence of MT was higher than that of ST (11.3% vs. 7.9%), with a prevalence ratio of 1.42 (*p*<0.001). In addition, exposure to bushes and living in sparsely populated areas are significant risk factors for MT. These findings indicate that MT is a neglected disease; hence, patients with suspected rickettsial infections, especially those with risk factors, should undergo comprehensive rickettsial infection testing, including MT testing.

**Article Summary Line:** This is a large epidemiological study in Japan to elucidate the seroprevalence and predictors of *Rickettsia typhi*, which may be neglected today.

## □Introduction□

Murine typhus (MT), a flea-borne rickettsiosis caused by *Rickettsia typhi*, is a ubiquitous but clinically less recognizable disease than scrub typhus (ST) and spotted fever group rickettsioses (1,2). MT symptoms include fever, headache, and rashes and may mimic those of viral or other rickettsial infections. Laboratory diagnostics, such as indirect immunoperoxidase (IP) or indirect immunofluorescence (IF) assays, are commercially unavailable (1–3). Therefore, the difficulty in diagnostic confirmation makes MT neglected. Severe MT cases leading to hospitalization or death have been reported, although many patients recover with appropriate antibiotics or spontaneously (4,5). Clarifying epidemiologic evidence of MT will help clinicians recognize the disease, have high suspicion, and provide early treatment.

MT is distributed globally and is endemic in warm urban or coastal regions, where rats or cats, the reservoirs of *R. typhi*, tend to inhabit; however, epidemiological characteristics often differ from region to region (1). Reports from industrialized countries, such as the US (6–8), Greece (9), and Spain, (10,11) have accumulated. Epidemiological studies from the US have indicated that MT is endemic not in urban areas but in suburban areas (6,8), potentially due to a new transmission cycle between fleas and reservoirs, such as opossums (12,13). Additionally, various risk factors for infection have been identified, including high population density (14,15), coastal areas (14,15), short distances from residences to the market, highways, lakes, and ponds (16–18). Accumulating specific and local evidence from each region is required to elucidate the complete picture of MT epidemiology.

In Japan, MT was endemic before the 1950s (19), but has not been notifiable, and only a few cases have been reported since then (20–22). There have been no seroepidemiological studies with standard diagnostic tests such as IP or IF. Consequently, the epidemiological characteristics remain unknown, potentially resulting in MT being unrecognized and neglected.

In this study, we aimed to estimate rickettsial seroprevalence, primarily MT, in a rickettsia-endemic area in Japan and to characterize the demographic and geographic informational risk factors for MT.

## □Methods□

### Study design and settings

This cross-sectional study was conducted in two local municipalities (Otaki and Katsuura, Japan) and Kameda Medical Center (Kamogawa, Japan) in the southern part of the Boso Peninsula, which is a predominantly mountainous agricultural region with a long coastline bordered by the Pacific Ocean and Tokyo Bay. According to the 2015 census, the populations of Otaki and Katsuura were 9,843 and 19,248, respectively (https://www.e-stat.go.jp/). Kameda Medical Center is a tertiary hospital with 865 acute beds that receives patients from all over the prefecture. We chose multiple sites for our study because rickettsioses, including ST and Japanese spotted fever (JSF) are endemic at these sites and are reported annually (23); epidemiological studies have been conducted previously (19); and we wanted to ensure the diversity of the participants. We followed the STROBE statement to report our study (24).

### Study participants

Participants who underwent health checkups at each site from August 2020 to November 2020 were registered. The health checkup services in Otaki and Katsuura were provided mainly to National Health Insurance subscribers, whereas those in Kameda Medical Center were provided for people who preferred, regardless of insurance type.

### Data collection and measurements

Questionnaires were distributed to participants during health checkups, and the following data were collected: medical history of rickettsioses, environmental exposure to mountains, agriculture, and bushes, and residential addresses of the participants. In addition, population density and land use area per 1 km square (coasts, forests, farmland, rivers/lakes, and wilderness), as registered in the 2015 national data (https://nlftp.mlit.go.jp/index.html), were linked to each participant’s address to obtain the population density, and the area of each land use within a 500-meter radius of the participant’s address. All of these data were collected using an open-source geographic information system (QGIS 3.16, https://qgis.org/ja/site/).

### Outcomes

The primary outcomes were MT seroprevalence and ratio of MT to ST seroprevalence. ST, caused by *Orientia tsutsugamushi*, was selected as the comparator outcome in this study, as ST is a notifiable and the most endemic rickettsial infection in Japan. Additionally, the seroprevalence of JSF caused by *Rickettsia japonica* was evaluated to address the possibility of an apparent excess seroprevalence of MT due to serological cross-reactivity in the genus *Rickettsia* (25,26). This allows for a potentially improved interpretation of the MT seroprevalence. It is noteworthy that *O. tsutsugamushi* is not serologically cross-reactive with *Rickettsia* spp., such as *R. typhi* and *R. japonica* (27). Upon the participants’ consent, serum rickettsial antibodies were measured using residual serum (0.5 mL) from blood samples collected for health checkups. The samples froze at −20 °C were sent to the Mahara Institute of Medical Acarology (Anan, Japan) for IP to measure IgG antibodies of six serotypes of *O. tsutsugamushi* (Kato, Karp, Gilliam, Irie/Kawasaki, Hirano/Kuroki, and Shimokoshi), *R. japonica* (Aoki strain), and *R. typhi* (Wilmington strain) (27,28). These samples were diluted from 1:40 to 1:40,960. For each antibody, seropositivity was defined as a ratio ≥1:40, and ST was defined as positive for any of the *O. tsutsugamushi* serotypes. We described the geographical distribution of *R. typhi* and *O. tsutsugamushi* IgG positives on the map using population density information depicted in QGIS 3.16.

### Statistical analyses

All statistical analyses were performed using Stata version 17.0 (StataCorp LLC, College Station, Texas, USA). Participant characteristics were summarized using medians and 5th and 95th percentiles for continuous variables, and numbers and percentages for categorical variables. The seroprevalence of MT and ST and primary outcome measures were estimated. As the seropositivity rates of MT and ST can be regarded as paired binomial data, the difference in their prevalence was tested using the McNemar test (29). To estimate the magnitude of the difference in seropositivity between the two different antibody assays (i.e., MT and ST) (30), conditional Poisson regression with a robust variance estimator was used (31,32). Furthermore, to test whether the magnitude of the ratios of prevalence differed across the study sites, an interaction between the pair and site variables was added to the regression model (30). The Wald test was used to test interactions. Finally, to explore the factors associated with MT seropositivity, a logistic regression model was fitted using clinical factors, environmental exposure, and geographic characteristics as explanatory variables. As there were missing values for population density and environmental exposure, multiple imputation (MI) was performed assuming that missing values occurred at random (33). For this study, five complete datasets were generated using MI with chained equations. Odds ratios obtained from the imputed data were combined according to Rubin’s rule. Two-sided *p* < 0.05 was considered statistically significant.

### Ethical approval

The study was approved by the Institutional Review Boards of Nagasaki University and Fukushima Medical University (no. 200305230-2, no. 2022–190). Written consent was obtained from all participants.

## □Results□

### Participants’ characteristics

Of the 2,382 participants (Otaki, 1,071; Kameda, 1,019; Katsuura, 292), 50.5% were women, with a median age of 67 years.

Participant characteristics and residential geographical features according to seropositivity for *R. typhi* IgG are shown in Table 1. The median population density of MT-positive residents was 244 persons/km^2^, whereas that of MT-negative residents was slightly higher than that of MT-positive residents (356 persons/km^2^). The median of coasts was recorded at 0 m^2^ in both the seropositive and seronegative groups, with the 5th to 95th percentiles showing 0 to 235,132 and 0 to 232,102, respectively. The participant characteristics and residential geographical features of *O. tsutsugamushi* seropositivity are presented in Appendix Table 1.

**Table 1.** Participants’ characteristics and their residential geographic features by seropositivity of *Rickettsia typhi* IgG*

|  | Murine typhus, Positive | Murine typhus, Negative | Total |
| --- | --- | --- | --- |
| Characteristic | (N = 269) | (N = 2,113) | (N = 2,382) |
| Female sex – no. (%) | 123 (45.7) | 1,080 (51.1) | 1,203 (50.5) |
| Age group, years – no. (%) |  |  |  |
| ≤40 | 2 (0.7) | 109 (5.2) | 111 (4.6) |
| 41–50 | 11 (4.1) | 269 (12.7) | 280 (11.8) |
| 51–60 | 15 (5.6) | 350 (16.6) | 365 (15.3) |
| 61–70 | 71 (26.4) | 695 (32.9) | 766 (32.2) |
| 71–80 | 113 (42) | 576 (27.2) | 689 (28.9) |
| ≥81 | 57 (21.2) | 114 (5.4) | 171 (7.2) |
| Site – no. (%) |  |  |  |
| Otaki | 180 (66.9) | 891 (42.2) | 1,071 (45) |
| Katsuura | 42 (15.6) | 250 (11.8) | 292 (12.2) |
| Kameda | 47 (17.5) | 972 (46) | 1,019 (42.8) |
| Past medical history – no. (%) |  |  |  |
| None | 227 (84.4) | 1,787 (85) | 2,014 (84.9) |
| Scrub typhus | 8 (3.0) | 32 (1.5) | 40 (1.7) |
| Japanese spotted fever | 2 (0.7) | 0 (0) | 2 (0.1) |
| Both | 0 (0) | 1 (0.1) | 1 (0) |
| Unknown | 32 (11.9) | 283 (13.5) | 315 (13.3) |
| Missing – no. | 0 | 10 | 10 |
| Environmental exposure – no. (%) |  |  |  |
| Mountains |  |  |  |
| Yes | 70 (26) | 456 (21.6) | 526 (22.1) |
| No | 199 (74) | 1,657 (78.4) | 1,856 (77.9) |
| Agriculture |  |  |  |
| Yes | 135 (50.2) | 766 (36.3) | 901 (37.8) |
| No | 134 (49.8) | 1,347 (63.7) | 1,481 (62.2) |
| Bushes |  |  |  |
| Yes | 141 (52.4) | 856 (40.5) | 997 (41.9) |
| No | 128 (47.6) | 1,257 (59.5) | 1,385 (58.1) |
| Population density |  |  |  |
| – persons/km<sup>2</sup> (5th–95th percentile) | 244 (32–1,207) | 356 (45–3,354) | 306 (44–3,148) |
| Missing – no. | 3 | 26 | 29 |
| Coasts |  |  |  |
| – m<sup>2</sup> (5th–95th percentile) | 0 (0–235,132) | 0 (0–232,102) | 0 (0–233,502) |
| Missing – no. | 0 | 3 | 3 |
| Forests |  |  |  |
| – m<sup>2</sup> (5th–95th percentile) | 307,376 (76,276–664,633) | 269,342 (6,023–616,031) | 273,757 (7,231–621,401) |
| Missing – no. | 0 | 3 | 3 |
| Farmland |  |  |  |
| – m<sup>2</sup> (5th–95th percentile) | 236,624 (14,833–458,114) | 233,350 (0–486,875) | 233,414 (0–483,051) |
| Missing – no. | 0 | 3 | 3 |
| Rivers and Lakes |  |  |  |
| – m<sup>2</sup> (5th–95th percentile) | 17,596 (0–101,103) | 13,917 (0–84,788) | 14,279 (0–86,124) |
| Missing – no. | 0 | 3 | 3 |
| Wilderness |  |  |  |
| – m<sup>2</sup> (5th–95th percentile) | 0 (0–27,181) | 0 (0–25,095) | 0 (0–25,391) |
| Missing – no. | 0 | 3 | 3 |

\*Continuous and categorical variable data are presented as median (5th–95th percentile) and frequency (%), respectively.

### Geographic distribution and the level of antibody titers in MT and ST seropositive participants

The residential locations of *R. typhi*-seropositive participants were distributed mainly in Otaki, Katsuura, and the entire southern part of the Boso Peninsula (Figure 1). The distribution map of the ST seropositive participants is shown in Appendix Figure 1.

**Figure 1.**
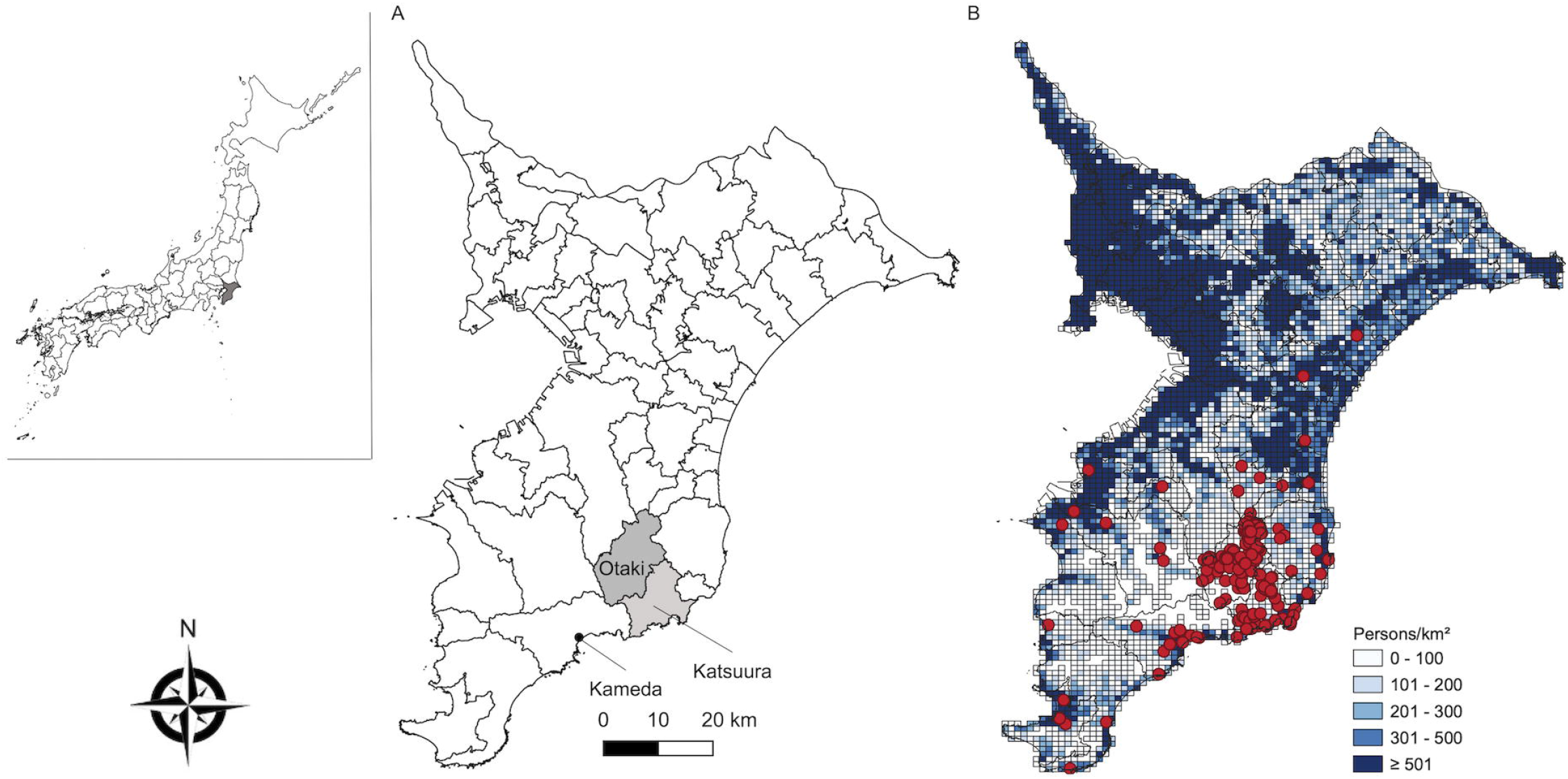
Geographical distribution of *Rickettsia typhi* IgG seropositive participants (A) This map illustrates the study sites (Otaki, Katsuura, and Kameda Medical Center), located in the southern part of the Boso Peninsula. (B) The living locations of *Rickettsia typhi* IgG seropositive individuals are indicated by red spots on the map containing population density data depicted by a white to dark blue gradient mesh.

Table 2 shows the distribution of antibody titers in *R. typhi* IgG-positive individuals, which is expressed as the reciprocal of the highest dilution of the serum that tested positive. Although approximately 60% of the participants had lower titers of ≤160, 20 participants had titers ≥1,280, and four participants’ titers reached ≥40,960. The counterparts of *O. tsutsugamushi* IgG-positive participants are described in Appendix Table 2.

**Table 2.** The distribution of antibody titers in *Rickettsia typhi* IgG-positive individuals*

| Antibody titers – no. (%) | Seropositive participants |
| --- | --- |
|  | (N = 269) |
| 40 | 53 (19.7) |
| 80 | 33 (12.3) |
| 160 | 80 (29.7) |
| 320 | 37 (13.7) |
| 640 | 46 (17.1) |
| 1280 | 5 (1.9) |
| 2560 | 8 (3.0) |
| 5120 | 2 (0.7) |
| 10240 | 1 (0.4) |
| 20480 | 0 (0) |
| ≥40960 | 4 (1.5) |

\*The titer indicates the reciprocal of the highest dilution of the serum, which is illustrated by the
blue or blue-black colored rickettsial particles.

### Analysis of the differences in seroprevalence between MT and ST

Overall, the seroprevalence of MT was 11.3% (95% confidence interval [CI] 10.0–12.6), which was higher than that of ST (7.9%, 95% CI 6.9–9.1) (*p*<0.001 by McNemar test, ratio of seropositivity 1.42, 95% CI 1.20–1.68) (Figure 2).

**Figure 2.**
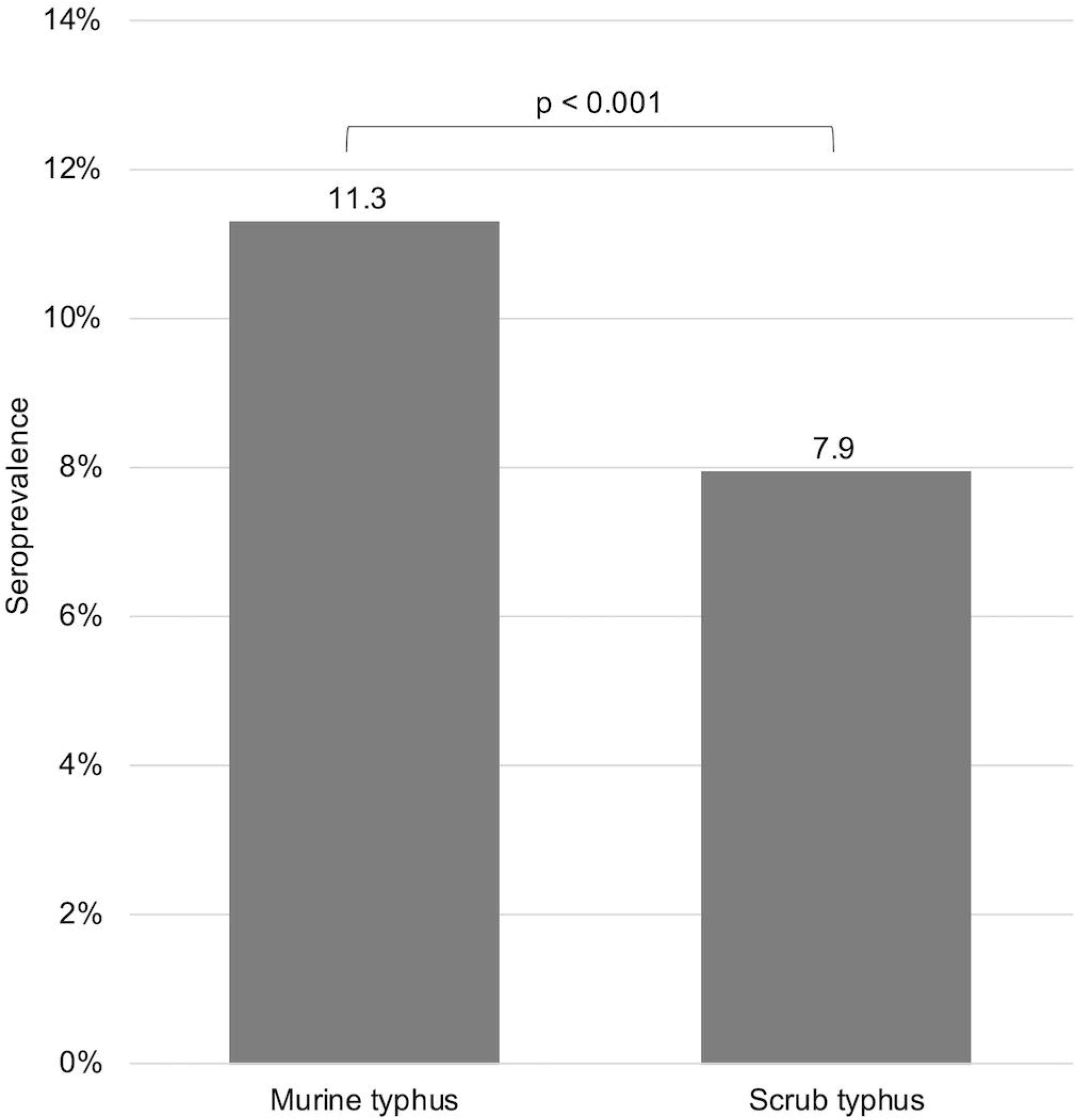
Seroprevalence of murine typhus and scrub typhus* *Murine typhus and scrub typhus represent 11.3% and 7.9% of all participants, respectively. The seroprevalence of both was compared using McNemar’s test. The ratio of seropositivity was estimated at 1.42, with a 95% confidence interval of 1.20–1.68.

The cross-reactivity of JSF with MT may result in a spurious increase in MT seroprevalence. Hence, JSF seroprevalence was examined and found to be positive for *R. japonica* IgG in 204 of 2,382 patients, with an estimate of 8.6% (95% CI 7.5–9.8), which was lower than that of MT.

The estimated prevalence ratios of MT to ST by the aforementioned three sites based on the conditional Poisson regression were 1.63 (95% CI, 1.32–2), 1.05 (95% CI 0.73–1.5), and 1.23 (95% CI, 0.79–1.92) in Otaki, Katsuura, and Kameda, respectively. However, the Wald test for the study-site difference in the seroprevalence ratio showed a *p*-value of 0.0932, indicating that the null hypothesis of no study-site difference in the seropositivity ratio between MT and ST could not be rejected.

### Predictors of MT seropositivity

The results of the multivariate analysis performed to explore the predictors of MT seropositivity are shown in Figure 3. The factors associated with MT seropositivity were age (per 10-year increase, adjusted odds ratio [aOR] 2.09, 95% CI 1.80–2.42), low population density (per 1000 persons/km^2^ increase, aOR 0.59, 95% CI 0.40–0.86), and history of bush exposure (aOR 1.39, 95% CI 1.01–1.92).

**Figure 3.**
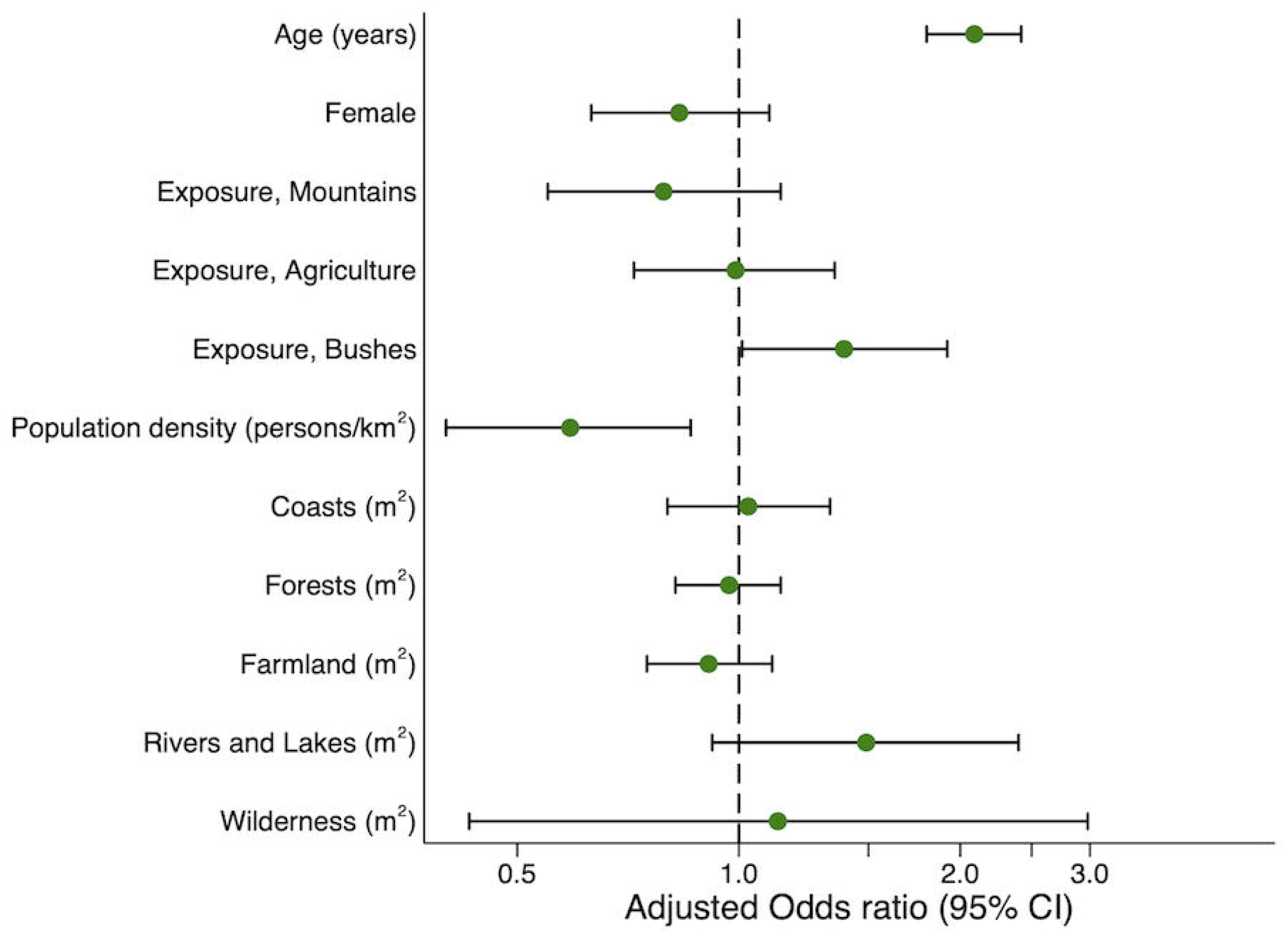
Predictors of *Rickettsia typhi* IgG seropositivity Adjusted odds ratios are shown for age per 10-year increase, population density per 1000 persons/km^2^ increase, and residential geographic features such as coasts, forests, farmland, rivers and lakes, and wilderness per 10 ha increase.

In contrast, the extent to which the coasts occupied a residential area (per 10 ha increase, aOR 1.03, 95% CI 0.80–1.33), forests (per 10 ha increase, aOR 0.97 [95% CI 0.82–1.14]), farmland (per 10 ha increase, aOR 0.91 [95% CI 0.75–1.11]), rivers and lakes (per 10 ha increase, aOR 1.49 [95% CI 0.92–2.40]), and wilderness (per 10 ha increase, aOR 1.13 [95% CI 0.43– 2.98]) were not associated with MT seropositivity. Other factors were also not associated with MT seropositivity: women (aOR 0.83 [95% CI 0.63–1.10]), history of mountain exposure (aOR 0.79 [95% CI 0.55–1.14]), and history of agricultural exposure (aOR 0.99 [95% CI 0.72–1.35]).

## □Discussion□

In this large seroepidemiological study of residents in a rickettsia-endemic area, where the incidence of ST has been most frequently reported in Japan, the seroprevalence of MT was much higher than that of ST among the general population. Furthermore, environmental exposure to bushes and low population density in residential areas were identified as predictors of MT seroprevalence, whereas residential geographic features, such as coasts, were not associated with MT seroprevalence. This is the first report on the epidemiological predominance of MT and its risk factors in Japan. Our study found a higher prevalence of MT than previous estimations, which urges clinicians to advance the diagnosis and treatment of rickettsiosis and its epidemiology.

The present study shows the predominance of MT while contrasting it with another epidemic rickettsiosis (ST), which is rarely reported elsewhere in the world in terms of the uniqueness of its analytical design. In addition, it quantitatively supports the findings of previous studies that suggest MT’s prevalence in Japan.

As the present study used the IP method, which is the current standard serological assay, the finding of the predominance of MT seroprevalence over ST, which is notifiable and still prevalent, is highly credible and surprising. In Japan, only six sporadic cases of MT between 1977 and 2013 and an outbreak in one prefecture in 1994 have been reported (20–22). In 1950, an endemic disease called twenty-day fever was prevalent in the study area, and its real cause was confirmed to be ST and MT by the Weil-Felix test (WFT) and culture (19). An epidemiological study of this disease was conducted to assess the epidemic status; however, its diagnostic method relied on the classic WFT (19). The WFT has limited diagnostic accuracy and can cause misclassification between rickettsioses (34,35); therefore, reliable seroprevalence of MT based on the assay cannot be obtained. Contrary to a previous study, we could differentiate IgG against *R. typhi* from that against *O. tsutsugamushi* using the standard diagnostic test, thereby confirming the estimation of the seroprevalence of MT and indicating its predominance.

In this study, the seroprevalence of MT (11%) was equal to or higher than that reported in developing countries. Thus, the present study illustrates that MT is a prevalent and possibly re-emerging infectious disease in developed countries. MT is ubiquitous in developing countries, with seroprevalence rates of 6.5% and 5.4% in northern Vietnam (36) and southern India (18), respectively. In addition, the incidence of MT has temporarily decreased, but has become a problem in developed countries such as the US (California and Texas) (6–8), Greece (9), and Spain (10,11). One possible reason for the increase in MT in California and Texas is the improved disease recognition by clinicians through local epidemiological studies (8,37). Another is a change in the transmission route from a rat flea (*Xenopsylla cheopis*)-rat transmission cycle to a cat flea (*Ctenocephalides felis*)-opossum or cat transmission cycle (13). In these areas, a new transmission cycle has been hypothesized based on local reports and studies (12,38). Thus, given the high seroprevalence of MT in Japan, case accumulation is crucial to clarify not only the disease characteristics and precise epidemiology, but also the possibility of a unique transmission cycle.

The observed increase in seroprevalence of MT with a decrease in residential population density shown in this study contradicts previous studies that showed that the urban environment is a risk factor for MT. In general, the reasons why MT cases are distributed mainly in coastal areas close to seaports and/or urban areas with high population densities are attributed to the introduction of infected rats and their fleas from ships and the environment conducive for rat proliferation (14,15). Indeed, the seroprevalence rate of *R. typhi* in urban rats was much higher than that in rural rats, and in Texas, 94% of the rats in surveyed urban facilities were seropositive (39). Rats from Taiwan’s most important international trading seaports had the highest seropositivity rates among those captured at airports and harbors (40).

While the rat-flea-rat cycle is the primary route of MT transmission, domestic cats and dogs, opossums and cat-fleas cause a new MT transmission cycle, especially in suburban areas with small populations (13,38,41). Since susceptibility to MT is believed to be associated more with vector and reservoir populations than with humans, more rats and their fleas or other vectors and reservoirs may be present for a greater MT-positivity rate in less densely populated areas, as observed in this study. Therefore, further studies are warranted to identify vectors and reservoirs in less-populated areas.

The differences in the types of risk factors between this study and previous studies may reflect the differences in the presence of factors related to contact with vectors and reservoirs at each study site. In this study, exposure to bushes was identified as a risk factor for MT seroprevalence, whereas residential environments, including coasts, rivers, and lakes, were not correlated. In contrast, in Vientiane, Laos, MT seroprevalence was associated with short distances between houses and markets (<750 m) and residences in areas with high building density (16); in Tanzania, short distances between houses and highways, and sparse vegetation (17); and in South India, pet dog ownership and the presence of lakes and ponds within 300 m (18). These studies have suggested that predictors of MT with possible links to vector and reservoir populations may vary by individual study sites, suggesting that the ecology of MT varies between study sites. Therefore, we believe that exploration of region-specific predictors is essential to clarify the full scope of MT epidemiology.

As this study confirms the predominance of MT, which is infrequently reported in Japan, a comprehensive examination for rickettsiosis, including MT, should be considered when patients are suspected of having these febrile illnesses in rickettsial endemic areas, especially those with a history of bush exposure or those living in sparsely populated areas. The importance of including MT in the differential list should not be underestimated. Many MT patients recover completely with or without antibiotics, while some may result in serious outcomes, such as hospitalization or death (4,5). However, diagnosing MT solely based on symptoms is very difficult, because MT presents with nonspecific symptoms such as fever, headache, and rash, which are also seen in other rickettsial infections and in some viral infections such as COVID-19 (3,5,42). As MT can present with intermediate fevers of 7–28 days in untreated cases (5,43), it may be reasonable to proactively test for MT in cases with these presentations and the epidemiologic characteristics.

Additionally, the present finding of the predominance of MT in regions where MT was believed to be almost non-existent demonstrates the possibility that an unexpectedly high seroprevalence of MT could be detected by performing epidemiological surveys of MT in previously unreported areas, including developed countries. Thus, MT may remain unrecognized, unless such studies have been conducted.

The present study has several strengths. First, the predominance of MT seroprevalence over ST in the general population of the study area was convincingly demonstrated by comparing the prevalence of MT with that of ST using paired binomial data. Second, the use of individual residential addresses and the QGIS geographic information system enabled a more precise quantification of the association of MT with population density and residential environment.

Instead of building density and distance from land use, we assessed the population density of the residential address and the surrounding land use surface area; thus, it is considered a more accurate representation of the residential neighborhood environment.

The present study has several limitations. First, MT seropositivity based on IgG titers may reflect a participant’s long-term pre-existing infection. Previous studies have shown that IgG titers for MT gradually decrease over time after infection (44). Thus, MT-positive cases without high IgG titers may not represent a particularly recent history of infection. However, 20 participants with MT antibody titers ≥1,280-fold may have had a recent infection. Second, IgG positivity to *R. typhi* could indicate cross-reactivity to *R. japonica* and represent a misclassification of JSF (25,26). However, our previous study showed that cross-reactivity to *R. typhi* was present in approximately 20% of patients with clinically and serologically confirmed JSF, indicating that the influence of cross-reactivity is limited (28). Furthermore, the seropositivity of MT in this study was 11.3%, which was higher than the 8.6% observed in the JSF. Collectively, these results suggest that only a portion of JSF causes MT cross-reactivity and that the higher seroprevalence of MT than JSF indicates a true positive for the majority of MT. Finally, since this study was conducted on health examination participants, it may not reflect the seroprevalence in the general population. However, we believe that our findings on MT seroprevalence are similar throughout the southern Boso Peninsula, as there were no differences at the site.

## Supporting information

STROBE_checklist

## Data Availability

All data produced in the present study are available upon reasonable request to the authors.

## □Conclusion□

The present study demonstrated that the seroprevalence of MT was higher than that of ST in rickettsial-endemic areas of Japan, indicating that MT is neglected. This study highlights the need to consider MT as a differential diagnosis, as well as other rickettsial infections, in patients presenting with clinical symptoms suggestive of rickettsial infections, intermediate fevers, and fevers of unknown origin. Therefore, a comprehensive examination of rickettsial infections, including MT testing, is essential, especially in patients living in sparsely populated areas, or who may have been exposed to bushes.

## Acknowledgments and Financial support

We thank the staff at municipal offices in Otaki and Katsuura and at Kameda Medical Center for collecting the questionnaires and blood samples. This work was supported by JSPS KAKENHI (grant number JP 19K23972).

## Author Bio

T. Aita is an internist specializing in general internal medicine and clinical epidemiology. He belongs to the Department of General Internal Medicine, Fukushima Medical University as a teaching/research associate, and is interested in research on the epidemiology of diseases, diagnostic accuracy, and infections, such as rickettsia.

## Appendix Table

Appendix Table Legends.

**Table 1.** Participants’ characteristics and their residential geographic features by *Orientia tsutsugamushi* IgG seropositivity*

| Characteristic | Scrub typhus, Positive<br>(N = 189) | Scrub typhus, Negative<br>(N = 2,193) | Total<br>(N = 2,382) |
| --- | --- | --- | --- |
| Female sex – no. (%) | 101 (53.4) | 1,102 (50.3) | 1,203 (50.5) |
| Age group, years – no. (%) |  |  |  |
| ≤40 | 5 (2.7) | 106 (4.8) | 111 (4.6) |
| 41–50 | 10 (5.3) | 270 (12.3) | 280 (11.8) |
| 51–60 | 18 (9.5) | 347 (15.8) | 365 (15.3) |
| 61–70 | 65 (34.4) | 701 (32.0) | 766 (32.2) |
| 71–80 | 66 (34.9) | 623 (28.4) | 689 (28.9) |
| ≥81 | 25 (13.2) | 146 (6.7) | 171 (7.2) |
| Site – no. (%) |  |  |  |
| Otaki | 111 (58.7) | 960 (43.8) | 1,071 (45.0) |
| Katsuura | 35 (18.5) | 257 (11.7) | 292 (12.2) |
| Kameda | 43 (22.8) | 976 (44.5) | 1,019 (42.8) |
| Past medical history – no. (%) |  |  |  |
| None | 137 (72.3) | 1,877 (85.9) | 2,014 (84.9) |
| Scrub typhus | 27 (14.4) | 13 (0.6) | 40 (1.7) |
| Japanese spotted fever | 1 (0.5) | 1 (0.05) | 2 (0.1) |
| Both | 0 (0) | 1 (0.05) | 1 (0) |
| Unknown | 22 (11.8) | 293 (13.4) | 315 (13.3) |
| Missing - no. | 2 | 8 | 10 |
| Environmental exposure – no. (%) |  |  |  |
| Mountains |  |  |  |
| Yes | 57 (30.2) | 469 (21.4) | 526 (22.1) |
| No | 132 (69.8) | 1,724 (78.6) | 1,856 (77.9) |
| Agriculture |  |  |  |
| Yes | 89 (47.1) | 812 (37.0) | 901 (37.8) |
| No | 100 (52.9) | 1,381 (63.0) | 1,481 (62.2) |
| Bushes |  |  |  |
| Yes | 91 (48.1) | 906 (41.3) | 997 (41.9) |
| No | 98 (51.9) | 1,287 (58.7) | 1,385 (58.1) |
| Population density |  |  |  |
| – persons/km<sup>2</sup> (5th–95th percentile) | 194 (31–1,207) | 335 (46–3,166) | 306 (44–3,148) |
| Missing – no. | 3 | 26 | 29 |
| Coasts |  |  |  |
| – m<sup>2</sup> (5th–95th percentile) | 0 (0–17,648) | 0 (0–237,384) | 0 (0–233,502) |
| Missing – no. | 0 | 3 | 3 |
| Forests |  |  |  |
| – m<sup>2</sup> (5th–95th percentile) | 341,158 (54,779–673,819) | 266,640 (6,272–619,733) | 273,757 (7,231–621,401) |
| Missing – no. | 0 | 3 | 3 |
| Farmland |  |  |  |
| – m<sup>2</sup> (5th–95th percentile) | 245,500 (22,140–462,920) | 233,091(0–483,615) | 233,414 (0–483,051) |
| Missing – no. | 0 | 3 | 3 |
| Rivers and Lakes |  |  |  |
| – m<sup>2</sup> (5th–95th percentile) | 13,583 (0–110,673) | 14,332 (0–85,033) | 14,279 (0–86,124) |
| Missing – no. | 0 | 3 | 3 |
| Wilderness |  |  |  |
| – m<sup>2</sup> (5th–95th percentile) | 0 (0–14,629) | 0 (0–25,989) | 0 (0–25,391) |
| Missing – no. | 0 | 3 | 3 |

\*Continuous and categorical variables are presented as median (5th–95th percentile) and frequency (%), respectively.

**Table 2.** The distribution of antibody titers in six serotypes of *Orientia tsutsugamushi* (Kato, Karp, Gilliam, Irie/Kawasaki, Hirano/Kuroki, and Shimokoshi) IgG-positive individuals*

| Antibody titers | Kato | Karp | Gilliam | Irie/Kawasaki | Hirano/Kuroki | Shimokoshi |
| --- | --- | --- | --- | --- | --- | --- |
| – no. (%) | (N = 38) | (N = 78) | (N = 84) | (N = 93) | (N = 71) | (N = 76) |
| 40 | 20 (52.6) | 33 (42.3) | 18 (21.4) | 18 (19.4) | 21 (29.6) | 32 (42.1) |
| 80 | 6 (15.8) | 14 (17.9) | 14 (16.7) | 4 (4.3) | 16 (22.5) | 16 (21.0) |
| 160 | 9 (23.7) | 19 (24.4) | 18 (21.4) | 9 (9.7) | 24 (33.8) | 21 (27.6) |
| 320 | 2 (5.3) | 6 (7.7) | 9 (10.7) | 15 (16.1) | 3 (4.2) | 5 (6.7) |
| 640 | 1 (2.6) | 4 (5.1) | 8 (9.5) | 18 (19.4) | 4 (5.6) | 0 (0) |
| 1280 | 0 (0) | 0 (0) | 9 (10.7) | 6 (6.4) | 0 (0) | 0 (0) |
| 2560 | 0 (0) | 1 (1.3) | 5 (6.0) | 7 (7.5) | 2 (2.8) | 1 (1.3) |
| 5120 | 0 (0) | 1 (1.3) | 0 (0) | 8 (8.6) | 0 (0) | 0 (0) |
| 10240 | 0 (0) | 0 (0) | 1 (1.2) | 3 (3.2) | 1 (1.4) | 1 (1.3) |
| 20480 | 0 (0) | 0 (0) | 2 (2.4) | 2 (2.2) | 0 (0) | 0 (0) |
| ≥40960 | 0 (0) | 0 (0) | 0 (0) | 3 (3.2) | 0 (0) | 0 (0) |

\*The titer indicates the reciprocal of the highest dilution of the serum, which is illustrated by the blue or blue-black colored rickettsial particles.

## Appendix Figure

Appendix figure Legend.

**Figure 1.**
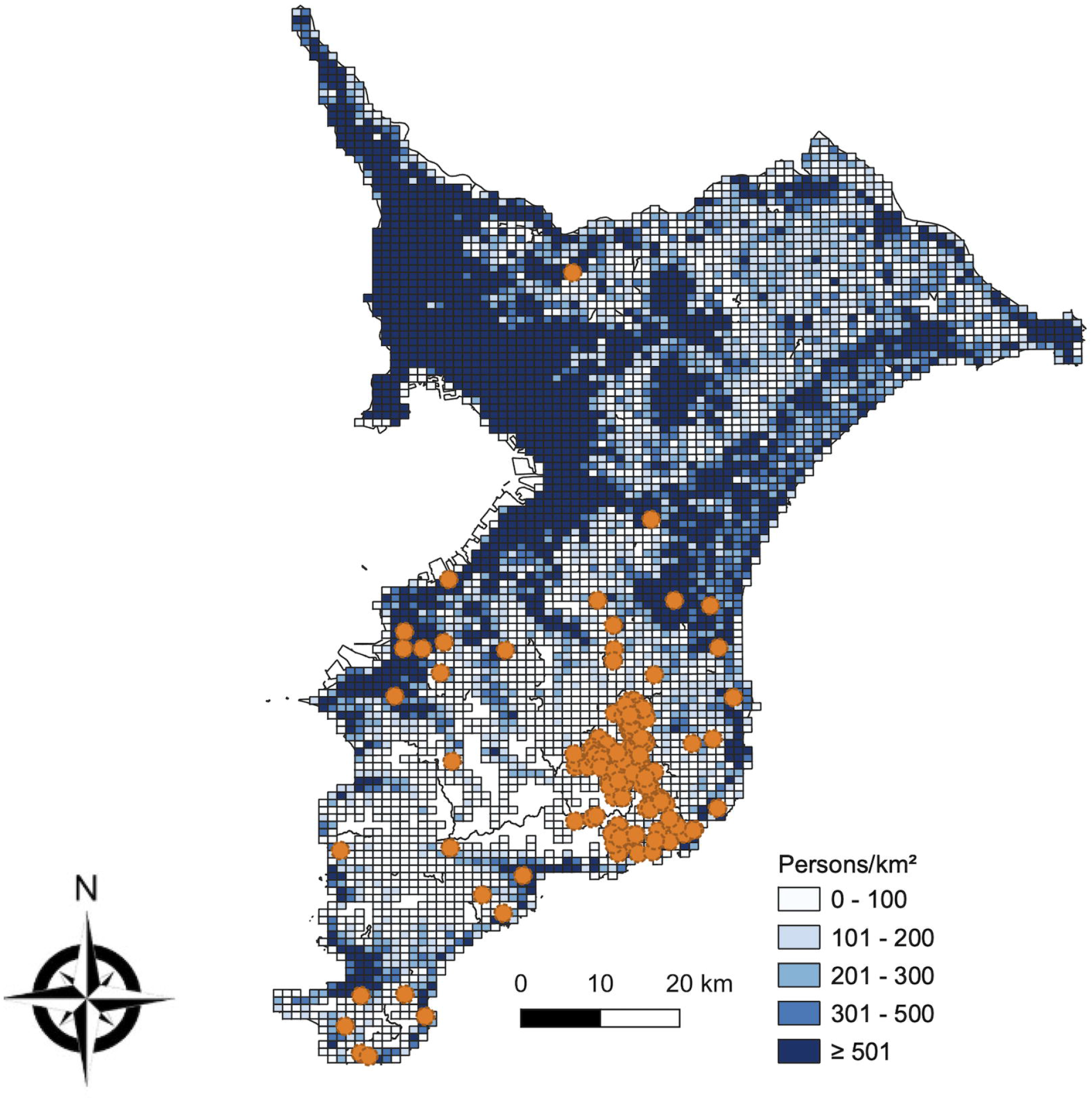
Geographical distribution in seropositive participants for *Orientia tsutsugamushi* IgG The locations where *Orientia tsutsugamushi* IgG-seropositive individuals lived are indicated by orange spots on the map containing population density data depicted by a white to dark blue gradient mesh.

## References

1. Civen R, Ngo V. Murine typhus: an unrecognized suburban vectorborne disease. Clin Infect Dis. 2008 Mar 15;46(6):913–8.

2. Blanton LS, Walker DH. Flea-Borne Rickettsioses and Rickettsiae. Am J Trop Med Hyg. 2017 Jan 11;96(1):53–6.

3. Faccini-Martínez ÁA, Walker DH, Blanton LS. Murine Typhus in Latin America: Perspectives of a Once Recognized but Now Neglected Vector-Borne Disease. Am J Trop Med Hyg [Internet]. 2022 Aug 15; Available from: http://dx.doi.org/10.4269/ajtmh.22-0070

4. Dumler JS, Taylor JP, Walker DH. Clinical and laboratory features of murine typhus in south Texas, 1980 through 1987. JAMA. 1991 Sep 11;266(10):1365–70.

5. Doppler JF, Newton PN. A systematic review of the untreated mortality of murine typhus. PLoS Negl Trop Dis. 2020 Sep;14(9):e0008641.

6. Murray KO, Evert N, Mayes B, Fonken E, Erickson T, Garcia MN, et al. Typhus Group Rickettsiosis, Texas, USA, 2003-2013. Emerg Infect Dis. 2017 Apr;23(4):645–8.

7. Centers for Disease Control and Prevention. Human flea-borne typhus cases in California. January 12, 2018 (https://www.cdph.ca.gov/Programs/CID/DCDC/CDPH%20Document%20Library/Flea-borneTyphusCaseCounts.pdf).

8. Ruiz K, Valcin R, Keiser P, Blanton LS. Rise in Murine Typhus in Galveston County, Texas, USA, 2018. Emerg Infect Dis. 2020 May;26(5):1044–6.

9. Labropoulou S, Charvalos E, Chatzipanagiotou S, Ioannidis A, Sylignakis P, Taka S, et al. Sunbathing, a possible risk factor of murine typhus infection in Greece. PLoS Negl Trop Dis. 2021 Mar;15(3):e0009186.

10. Rodríguez-Alonso B, Almeida H, Alonso-Sardón M, Velasco-Tirado V, Robaina Bordón JM, Carranza Rodríguez C, et al. Murine typhus. How does it affect us in the 21st century? The epidemiology of inpatients in Spain (1997-2015). Int J Infect Dis. 2020 Jul;96:165–71.

11. Robaina-Bordón JM, Carranza-Rodríguez C, Hernández-Cabrera M, Bolaños-Rivero M, Pisos-Álamo E, Jaén-Sánchez N, et al. Murine Typhus in Canary Islands, Spain, 1999-2015. Emerg Infect Dis. 2021 Feb;27(2):570–3.

12. Adams WH, Emmons RW, Brooks JE. The changing ecology of murine (endemic) typhus in Southern California. Am J Trop Med Hyg. 1970 Mar;19(2):311–8.

13. Blanton LS, Idowu BM, Tatsch TN, Henderson JM, Bouyer DH, Walker DH. Opossums and Cat Fleas: New Insights in the Ecology of Murine Typhus in Galveston, Texas. Am J Trop Med Hyg. 2016 Aug 3;95(2):457–61.

14. Traub R, Wisseman CL. The ecology of murine typhus-a critical review. Trop Dis Bull. 1978 Apr;75(4):237–317.

15. Azad AF. Epidemiology of murine typhus. Annu Rev Entomol. 1990;35:553–69.

16. Vallée J, Thaojaikong T, Moore CE, Phetsouvanh R, Richards AL, Souris M, et al. Contrasting spatial distribution and risk factors for past infection with scrub typhus and murine typhus in Vientiane City, Lao PDR. PLoS Negl Trop Dis. 2010 Dec 7;4(12):e909.

17. Dill T, Dobler G, Saathoff E, Clowes P, Kroidl I, Ntinginya E, et al. High seroprevalence for typhus group rickettsiae, southwestern Tanzania. Emerg Infect Dis. 2013 Feb;19(2):317–20.

18. Devamani CS, Schmidt WP, Ariyoshi K, Anitha A, Kalaimani S, Prakash JAJ. Risk factors for scrub typhus, murine typhus, and spotted fever seropositivity in urban areas, rural plains, and peri-forest hill villages in South India: A cross-sectional study. Am J Trop Med Hyg. 2020;103(1):238–48.

19. Tamiya T. Recent Advances in Studies of Tsutsugamushi Disease in Japan. 1962.

20. Takagi K, Iwasaki H, Kishi S, Nakamura T, Takada N, Ueda T. [Murine typhus infected in Oku-etsu area, Fukui Prefecture]. Kansenshogaku Zasshi. 2001 Apr;75(4):341–4.

21. Sakaguchi S, Sato I, Muguruma H, Kawano H, Kusuhara Y, Yano S, et al. Reemerging murine typhus, Japan. Emerg Infect Dis. 2004 May;10(5):964–5.

22. Infectious Agents Surveillance Report (IASR). 2013 Oct;34: 313–4. (in Japanese) [Internet] [Internet]. Available from: https://www.niid.go.jp/niid/ja/diseases/ra/rickettsia.html

23. Sando E, Suzuki M, Katoh S, Fujita H. Distinguishing Japanese Spotted Fever and Scrub Typhus. Emerg Infect Dis. 2018;24(9):1633–41.

24. von Elm E, Altman DG, Egger M, Pocock SJ, Gøtzsche PC, Vandenbroucke JP, et al. The Strengthening the Reporting of Observational Studies in Epidemiology (STROBE) statement: guidelines for reporting observational studies. J Clin Epidemiol. 2008 Apr;61(4):344–9.

25. Uchiyama T, Zhao L, Yan Y, Uchida T. Cross-reactivity of Rickettsia japonica and Rickettsia typhi demonstrated by immunofluorescence and Western immunoblotting. Microbiol Immunol. 1995;39(12):951–7.

26. Robinson MT, Satjanadumrong J, Hughes T, Stenos J, Blacksell SD. Diagnosis of spotted fever group Rickettsia infections: the Asian perspective. Epidemiol Infect. 2019 Oct 7;147:e286.

27. Sando E, Ariyoshi K, Fujita H. Serological Cross-Reactivity among Orientia tsutsugamushi Serotypes but Not with Rickettsia japonica in Japan. Tropical Medicine and Infectious Disease. 2018;3(3):74.

28. Aita T, Sando E, Katoh S, Hamaguchi S, Fujita H, Kurita N. Serological Cross-Reactivity Between Spotted Fever and Typhus Groups of Rickettsia Infection in Japan [Internet]. medRxiv. 2022 [cited 2022 Oct 18]. p. 2022.10.16.22281137. Available from: https://www.medrxiv.org/content/10.1101/2022.10.16.22281137v1.abstract

29. Pembury Smith MQR, Ruxton GD. Effective use of the McNemar test. Behav Ecol Sociobiol. 2020 Oct 10;74(11):133.

30. Cummings P, McKnight B, Greenland S. Matched cohort methods for injury research. Epidemiol Rev. 2003;25:43–50.

31. Cummings P, McKnight B. Analysis of Matched Cohort Data. Stata J. 2004 Aug 1;4(3):274–81.

32. Cummings P. Estimating Adjusted Risk Ratios for Matched and Unmatched Data: An Update. Stata J. 2011 Jul 1;11(2):290–8.

33. Donders ART, van der Heijden GJMG, Stijnen T, Moons KGM. Review: a gentle introduction to imputation of missing values. J Clin Epidemiol. 2006 Oct;59(10):1087–91.

34. La Scola B, Raoult D. Laboratory diagnosis of rickettsioses: current approaches to diagnosis of old and new rickettsial diseases. J Clin Microbiol. 1997 Nov;35(11):2715–27.

35. Stewart AG, Stewart AGA. An Update on the Laboratory Diagnosis of Rickettsia spp. Infection. Pathogens [Internet]. 2021 Oct 13;10(10). Available from: http://dx.doi.org/10.3390/pathogens10101319

36. Vu Trung N, Hoi LT, Thuong NTH, Toan TK, Huong TTK, Hoa TM, et al. Seroprevalence of scrub typhus, typhus, and spotted fever among rural and urban populations of northern Vietnam. Am J Trop Med Hyg. 2017;96(5):1084–7.

37. Erickson T, da Silva J, Nolan MS, Marquez L, Munoz FM, Murray KO. Newly Recognized Pediatric Cases of Typhus Group Rickettsiosis, Houston, Texas, USA. Emerg Infect Dis. 2017 Dec;23(12):2068–71.

38. Sorvillo FJ, Gondo B, Emmons R, Ryan P, Waterman SH, Tilzer A, et al. A suburban focus of endemic typhus in Los Angeles County: association with seropositive domestic cats and opossums. Am J Trop Med Hyg. 1993 Feb;48(2):269–73.

39. Irons JV, Murphy JN Jr, Davis DE. The distribution of endemic typhus in rats in Lavaca County, Texas. Public Health Rep. 1948 May 21;63(21):692–4.

40. Chiang PS, Su SW, Yang SL, Shu PY, Lee WP, Li SY, et al. Delayed correlation between the incidence rate of indigenous murine typhus in humans and the seropositive rate of Rickettsia typhi infection in small mammals in Taiwan from 2007-2019. PLoS Negl Trop Dis. 2022 Apr;16(4):e0010394.

41. Nogueras MM, Pons I, Pla J, Ortuño A, Miret J, Sanfeliu I, et al. The role of dogs in the eco-epidemiology of Rickettsia typhi, etiological agent of Murine typhus. Vet Microbiol. 2013 Apr 12;163(1–2):97–102.

42. Patel HM. Murine typhus mistaken for COVID-19 in a young man. BMJ Case Rep [Internet]. 2020 Nov 3;13(11). Available from: http://dx.doi.org/10.1136/bcr-2020-239471

43. Tsioutis C, Zafeiri M, Avramopoulos A, Prousali E, Miligkos M, Karageorgos SA. Clinical and laboratory characteristics, epidemiology, and outcomes of murine typhus: A systematic review. Acta Trop. 2017 Feb;166:16–24.

44. Halle S, Dasch GA. Use of a sensitive microplate enzyme-linked immunosorbent assay in a retrospective serological analysis of a laboratory population at risk to infection with typhus group rickettsiae. J Clin Microbiol. 1980 Sep;12(3):343–50.

